# Forward-looking serial intervals correctly link epidemic growth to reproduction numbers

**DOI:** 10.1101/2020.06.04.20122713

**Authors:** Sang Woo Park, Kaiyuan Sun, David Champredon, Michael Li, Benjamin M. Bolker, David J. D. Earn, Joshua S. Weitz, Bryan T. Grenfell, Jonathan Dushoff

## Abstract

The reproduction number ℛ and the growth rate *r* are critical epidemiological quantities. They are linked by generation intervals, the time between infection and onward transmission. Because generation intervals are difficult to observe, epidemiologists often substitute serial intervals, the time between symptom onset in successive links in a transmission chain. Recent studies suggest that such substitution biases estimates of ℛ based on *r*. Here we explore how these intervals vary over the course of an epidemic, and the implications for ℛ estimation. *Forward-looking* serial intervals, measuring time forward from symptom onset of an infector, correctly describe the renewal process of symptomatic cases and therefore reliably link ℛ with *r*. In contrast, *backward-looking* intervals, which measure time backward, and *intrinsic* intervals, which neglect population-level dynamics, give incorrect ℛ estimates. Forward-looking intervals are affected both by epidemic dynamics and by censoring, changing in complex ways over the course of an epidemic. We present a heuristic method for addressing biases that arise from neglecting changes in serial intervals. We apply the method to early (21 January – 8 February 2020) serial-interval-based estimates of ℛ for the COVID-19 outbreak in China outside Hubei province; using improperly defined serial intervals in this context biases estimates of initial ℛ by up to a factor of 2.6. This study demonstrates the importance of early contact-tracing efforts and provides a framework for reassessing generation intervals, serial intervals, and ℛ estimates for COVID-19.

**Significance Statement:** The generation- and serial-interval distributions are key, but different, quantities in outbreak analyses. Recent theoretical studies suggest that the two distributions give different estimates of the reproduction number ℛ as inferred from the observed exponential growth rate *r*. Here, we show that estimating ℛ based on *r* and the serial-interval distribution, when defined from the correct reference time and cohort, gives the same estimate as using *r* and the generation-interval distribution. We apply our framework to serial-interval data from the COVID-19 outbreak in China, outside Hubei province (January 21–February 8, 2020), revealing systematic biases in prior inference methods. Our study provides the theoretical basis for practical changes to the principled use of serial interval distributions in estimating ℛ during epidemics.

## 1 Introduction

The reproduction number ℛ is one of the most important characteristics of an emerging epidemic, such as the current pandemic of coronavirus disease 2019 (COVID-19) (Majumder and Mandl, 2020). The reproduction number is defined as the average number of secondary cases caused by a primary case. The value in a fully susceptible population — the “basic” reproduction number ℛ_0_— allows us to predict the extent to which an infection will spread in the population, and the amount of intervention necessary to eliminate it in simple cases (Anderson and May, 1991). Since the reproduction number represents an average (Diekmann et al., 1990; Anderson and May, 1991), it fails to capture heterogeneity among individuals or across space. The reproduction number also fails to provide any information about the time scale of disease transmission.

Estimating the reproduction number ℛ is often challenging. Direct estimates based on observed infections will typically be biased down when some infections cannot be observed. A common method of estimating ℛ near the beginning of an epidemic is based on the population-level exponential growth rate *r*, which can often be estimated robustly from case reports (Mills et al., 2004; Ma et al., 2014). The growth rate *r* and the reproduction number ℛ are linked by the generation-interval distribution Wallinga and Lipsitch (2007), where the generation interval is defined as the time between when an individual (infector) is infected and when that individual infects another person (infectee) (Svensson, 2007).

Since generation intervals measure time between infection events, which can be difficult to observe in practice, generation intervals are often replaced with serial intervals. The serial interval is defined as the time between when an infector and an infectee develop symptoms (Svensson, 2007). While generation and serial intervals both measure the time scale of disease transmission, they measure fundamentally different quantities. In particular, previous studies have noted that, in many contexts, serial intervals are expected to have larger variances than generation intervals but have the same mean in many contexts (Svensson, 2007; Klinkenberg and Nishiura, 2011; te Beest et al., 2013; Champredon et al., 2018). Serial intervals can in some cases even take negative values in the presence of presymptomatic transmission (He et al., 2020), whereas generation intervals must be positive.

Although these distributions were clearly and distinctly defined over a decade ago (Svensson, 2007), the need for a better conceptual and theoretical framework for understanding their differences is becoming clearer as the COVID-19 pandemic unfolds. Researchers continue to base inferences about COVID-19 on both generation and serial intervals without clearly distinguishing between them (e.g., Abbott et al. (2020); Du et al. (2020); He et al. (2020); Wu et al. (2020); Zhao et al. (2020)), and, in some cases, explicitly conflate the definitions of the two intervals (e.g., Anderson et al. (2020); Hellewell et al. (2020)). This confusion is apparent even in standard software for estimating ℛ, such as `EpiEstim`, in which the serial-interval distribution is used to infer time-dependent ℛ (Thompson et al., 2019). These studies are examples of many—indeed, it is a common practice to use the serial and generation intervals interchangeably.

One source of confusion arises from an apparent discrepancy between the generation-interval and serial-interval viewpoints. While the epidemic is growing exponentially, the spread of infection can be characterized as a *renewal process* based on previous incidence of infection, the associated generation-interval distribution, and the average infectiousness of an infected individual. It is well established that this renewal formulation allows us to link the exponential growth rate of an epidemic *r* with its reproduction number ℛ using the generation-interval distribution (Wallinga and Lipsitch, 2007). However, the serial-interval distribution also describes a renewal process — in this case, the creation of a new *symptomatic* case based on a symptomatic case in the previous generation. Since both renewal processes, based on either generation- or serial-interval distributions, describe the same underlying exponentially growing system, both should provide the same correct link between the reproduction number ℛ and the epidemic growth rate *r*.

In contexts where the serial- and generation-interval distributions differ, current theory has no explanation for how two different distributions could provide identical estimates of ℛ from *r*. In fact, recent theory suggest that using the serial-interval can underestimate the reproduction number (Britton and Scalia Tomba, 2019; Ganyani et al., 2020). However, these studies rely on *intrinsic* distributions of incubation periods and generation intervals that neglect the population-level dynamics of disease spread.

Here we show that, by correctly defining and calculating the “forward” serial-interval distribution (i.e., a distribution of serial intervals from a cohort of infectors that developed symptoms at the same time) that connects symptom onset dates, we can resolve this discrepancy. These forward intervals are different from the “intrinsic” serial intervals that previous studies have relied on (Svensson, 2007; Klinkenberg and Nishiura, 2011; te Beest et al., 2013; Champredon et al., 2018; Britton and Scalia Tomba, 2019). During an ongoing epidemic, all observed epidemiological delays (e.g., incubation period) between primary (e.g., infection) and secondary (e.g., symptom onset) events are subject to backward biases: when the incidence of primary events is increasing (or decreasing), we are more likely to observe shorter (respectively longer) intervals. In particular, when we consider forward serial-interval distributions, the incubation periods of the infectors are subject to backward biases because we have to look backward in time from their symptom onset to infection. Therefore, the realized incubation period distributions of the infector and the infectee can differ dynamically, even if the intrinsic analogues of the same distributions are expected to be equivalent.

We develop a cohort-based framework for characterizing and comparing realized serial intervals, as well as any other epidemiological delays, and show that the initial forward serial-interval distribution correctly estimates ℛ from *r*. Conversely, using inaccurately defined serial intervals or failing to account for changes in the observed serial-interval distributions over the course of an epidemic can considerably bias estimates of ℛ. For example, in our analysis of the COVID-19 serial intervals from China, outside Hubei province, we find that the original ℛ_0_ estimates based on aggregated serial-interval data underestimated ℛ_0_ by a factor of 2.0–2.6. We further lay out several principles to consider in using information about serial intervals and other epidemiological time delays to correctly infer the initial reproduction number during the early stages of an outbreak.

## 2 Methods

### 2.1 Intrinsic, forward, and backward delay distributions

A time delay between two epidemiological events can involve either one infected individual (e.g., incubation period: infection and symptom onset of an individual) or two — an infector and an infectee (e.g., generation and serial intervals). We define the delay as the time difference between the *primary* event and the *secondary* event. In some cases, the primary event always occurs before the secondary event (e.g., the time from infection to onset of symptoms in a single individual, or the generation interval between two individuals). In other cases, the delay can sometimes be negative (e.g., the time from onset of infectiousness to onset of symptoms in a single individual, or the serial interval between two individuals).

At the individual level, we can define the time distribution between a primary and a secondary event that we expect to observe for a single infected individual by averaging across individual characteristics — we refer to this distribution as the *intrinsic distribution*. For example, the intrinsic incubation period distribution describes the expected time distribution from infection to symptom onset of an infected individual. Likewise, the intrinsic generation-interval distribution describes the expected time distribution of infectious contacts made by an infected individual. However, the intrinsic time distributions are not always equivalent to the corresponding realized time distributions at the population level (i.e., the distribution of time between actual primary and secondary events that occur during an epidemic; see Fig. 1). For example, an infectious contact results in infection only if the contacted individual is susceptible (and has not already been infected) — this is one mechanism that causes realized generation intervals (time between actual infection events) to differ from from the intrinsic generation intervals (time between infection and infectious contacts) (Park et al., 2020). In this example, the difference between intrinsic and realized time distributions can be attributed to the fact that the fraction of susceptible individuals is itself dynamic.

**Figure 1:**
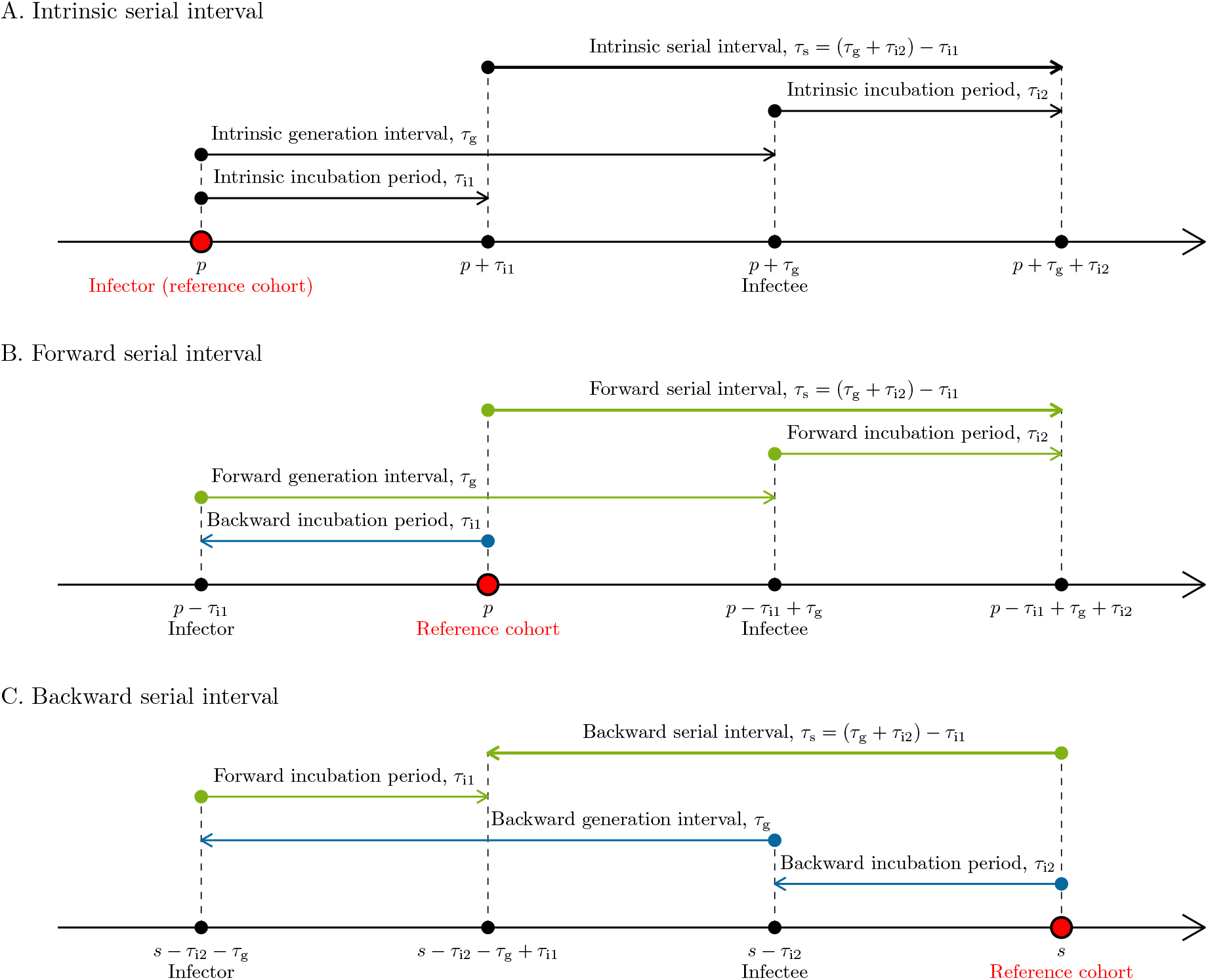
Illustration of intrinsic, forward and backward serial intervals. (A) The intrinsic serial interval for a cohort of individuals infected at time *p*. In this case, *τ*_i1_ is drawn from the intrinsic incubation period distribution; *τ*_g_ is drawn from the intrinsic generation-interval distribution; and *τ*_i2_ is drawn from the intrinsic incubation period distribution. (B) The forward serial interval for a cohort of infectors who became symptomatic at time *p*. In this case, *τ*_i1_ is drawn from the backward incubation period distribution; *τ*_g_ is drawn from the forward generation-interval distribution; and *τ*_i2_ is drawn from the forward incubation period distribution. (C) The backward serial interval for a cohort of infectees who became symptomatic at time *s*. In this case, *τ*_i1_ is drawn from the forward incubation period distribution; *τ*_g_ is drawn from the backward generation-interval distribution; and *τ*_i2_ is drawn from the backward incubation period distribution. Intrinsic intervals (black) reflect average of individual characteristics and are not dependent on population-level dynamics. Forward intervals (green) can change due to epidemiological dynamics (e.g., contraction of generation intervals through susceptible depletion). Backward intervals (blue) can change due to changes in cohort sizes even when forward intervals remain time-invariant.

At the population level, we model realized time delays between a primary and a secondary event from a cohort perspective. A cohort consists of all individuals whose (primary or secondary) event occurred at a given time. For example, when we are measuring incubation periods, a primary cohort consists of all individuals who became infected at time *p*, while a secondary cohort consists of all individuals whose symptom onset occurred at time *s*. Similarly, when we are measuring serial intervals, a primary cohort consists of all infectors who became symptomatic at time *p*. Then, for a primary cohort at time *p*, we can define the distribution of realized delays between primary and secondary events. We refer to this distribution as the forward delay distribution and denote it as *f*_*p*_(*τ*).

Likewise, we define the backward delay distribution *b*_*s*_(*τ*) for a secondary cohort at time *s*: The backward delay distribution describes the time delays between a primary and secondary events given that the secondary event occurred at time *s*. For example, the backward incubation period distribution at time *s* describes incubation periods for a *cohort* of individuals who became symptomatic at time *s*. Likewise, the backward serial-interval distribution at time *s* describes serial intervals for a *cohort* of infectees who became symptomatic at time *s*.

Both forward and backward perspectives must yield identical *measurement* (e.g., the length of the incubation period of a given individual is the same whether measured forward from the time of infection or backward from the time of symptom onset). Consequently, no matter how delays are distributed, if 𝒫 and 𝒮 represent the sizes of primary and secondary cohorts then we can express the total density of intervals *τ* between calendar time *p* and *s* (i.e., *τ* = *s* − *p*) as follows:

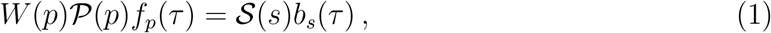

where *W* (*p*), the “weight” of the primary cohort, represents the average number of forward intervals that an individual in cohort 𝒫 (*p*) produces over the course of their infection. When we measure within-individual delays, we expect *W* (*p*) ≤ 1 because only a subset of individuals who experience the primary event (e.g., infection) will eventually experience the secondary event (e.g., symptom onset). For between-individual delays, we expect *W* (*p*) to change throughout an epidemic, because individuals infected earlier in an epidemic will infect more individuals on average than those infected later.

Substituting *p* = *s* − *τ*, it follows that

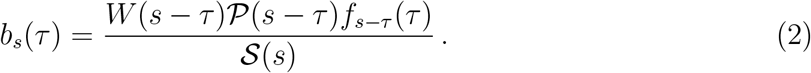

If we are considering incubation periods, the left hand side of this equation is the probability density that an individual who became symptomatic at time *s* had an incubation period of length *τ*. From the right hand side, we see that this probability density depends on the weight parameter *W* (*s* − *τ*) (in this case, the proportion of symptomatic infection), the time-varying primary cohort size at the earlier time 𝒫 (*s* − *τ*) (in this case, the number of individuals infected at time *s − τ*), and the forward delay distribution *f*_*s*−*τ*_ (*τ*) (in this case, the probability density that an incubation period that starts at time *s* − *τ* ends at time *s*).

Several different mechanisms drive the changes in forward and backward delay distributions over time. Typically, within-individual forward delay distributions are not directly affected by epidemic dynamics. Some realized forward distributions, like incubation period distributions, are equivalent to their intrinsic distributions and remain invariant at the time scale of an outbreak. Other realized distributions, like the distribution of time from symptom onset to testing, may change over the course of an epidemic due to changes in public-health policies or individual behavior. Between-individual forward delay distributions, such as generation- or serial-interval distributions, depend on epidemic dynamics. For example, forward generation intervals often become shorter as an epidemic progresses due to the dynamical process of susceptible depletion, as well as due to other factors like behavioral change or interventions (Kenah et al., 2008; Nishiura, 2010; Champredon and Dushoff, 2015): if it is harder to infect later in the course of infection, then proportionally more intervals will be short.

Eq. (2) suggests that backward delay distributions change over time even if their corresponding forward delay distribution does not change. Backward delay distributions depend on changes in the primary cohort size over time due to conditionality of observations: Conditioning on individuals whose secondary events have occurred at the same time means that we tend to observe shorter (or longer) inter-event delays when cohort size has been increasing (decreasing) through time. When incidence is growing exponentially, we can calculate the amount of bias exactly. Assuming that the forward delay distribution (*f*_*p*_(*τ*) ≈ *f*_0_(*τ*)) and the weight parameter (*W* (*p*) ≈ *W* (0)) remain constant during the exponential growth phase, we can substitute 𝒫 (*t*) = 𝒫 (0) exp(*rt*) in Eq. (2) to obtain:

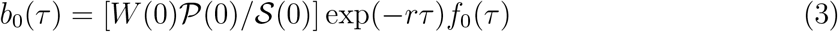

where *r* is the exponential growth rate. Since *b*_0_ is a probability distribution, 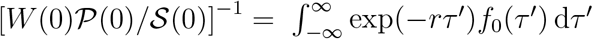 corresponds to the normalization constant. Therefore, the backward delay distribution during the exponential growth phase depends only on the exponential growth rate *r* and the initial forward delay distribution *f*_0_.

The mean backward interval will be always shorter than the mean forward interval as long as *r >* 0. Even for different epidemics of the same disease, we expect to observe shorter backward intervals within a fast-growing epidemic (high *r*), all else being equal. In general, the backward delay distribution will differ from the forward delay distribution (unless the disease is at equilibrium), even if we are measuring time delays that are intrinsic to the life history of a disease (e.g., the incubation period). These ideas apply to all epidemiological delay distributions and generalize the work by Champredon and Dushoff (2015) who compared forward and backward generation-interval distributions to describe realized generation intervals from the perspective of an infector and an infectee, respectively, as well as the work by Britton and Scalia Tomba (2019) who showed that Eq. (3) holds for the backward generation-interval distribution.

### 2.2 Realized serial-interval distributions

The serial interval is defined as the time between when an infector becomes symptomatic and when their infectee becomes symptomatic (Svensson, 2007). Previous studies have often expressed serial intervals *τ*_s_ in the form (Fig. 1A):

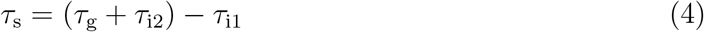

where *τ*_i1_ and *τ*_i2_ represent incubation periods of an infector and an infectee, respectively, and *τ*_g_ represents the generation interval between the infector and the infectee. These studies concluded that the serial and generation intervals have the same mean when *τ*_i1_ and *τ*_i2_ are drawn from the same distributions (Svensson, 2007; Klinkenberg and Nishiura, 2011; Champredon et al., 2018; Britton and Scalia Tomba, 2019). However, distributions of realized incubation periods, *τ*_i1_ and *τ*_i2_ will be identical only if we assume that they are intrinsic to individuals (and not dependent on epidemic dynamics at the population-level) — something that is generally true of forward but not backward incubation-period distributions. We refer to the definition Eq. (4) as the intrinsic serial interval (Fig. 1A).

To correctly link the realized serial-interval distribution to the renewal process between cases based on symptom onset dates, we must use the forward serial interval (i.e., use the perspective of a cohort of infectors that share the same symptom onset time). Given that an infector became symptomatic at time *p*, to calculate the forward serial interval we first go *backward* in time to when the infector was infected, and then forward in time to when the infectee was infected, and then forward again to when the infectee became symptomatic. In Fig. 1B, we see that *τ*_i1_ is drawn from the backward incubation period distribution of the cohort of infectors who became symptomatic at time *p*; *τ*_g_ is drawn from the forward generation-interval distribution of the cohort of infectors who became infected at time *p*−*τ*_i1_; and *τ*_i2_ is drawn from the forward incubation period distribution of the cohort of infectees who became infected at time *p* − *τ*_i1_ + *τ*_g_. Likewise, we can define the backward serial-interval distribution for a cohort of infectees who became symptomatic at time *s*(Fig. 1C). This conceptual framework demonstrates that the distributions of *τ*_i1_, *τ*_g_, and *τ*_i2_ (and therefore the distributions of realized serial intervals) depend on the reference cohort, which is defined by temporal direction (forward or backward) and a particular reference time.

To calculate realized serial-interval distributions, we begin by modeling 𝒯 (*p, s*): the total density of serial intervals that start (when infectors develop symptoms) at time *p* and end (when infectees develop symptoms) at time *s*. For simplicity, we assume that all infected individuals eventually develop symptoms. Then, the density of serial intervals between time *p* and *s*, given that the infectors became infected at time *α*_1_ ≤ *p* and the infectees became infected at time *α*_2_ ≤ *s*, depends on the amount of infection that occurs between time *α*_1_ and *α*_2_ as well as the density of forward incubation periods between *α*_1_ and *p* (realized incubation periods of infectors) and between *α*_2_ and *s*(realized incubation periods of infectees):

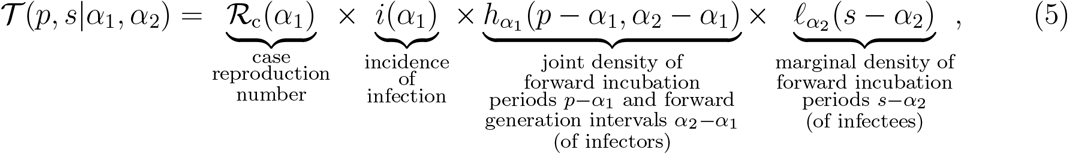

where the case reproduction number ℛ_c_(*α*_1_) is defined as the average number of secondary cases that a primary case infected at time *α*_1_ will generate over the course of their infection (Fraser, 2007). We describe the forward incubation periods and the forward generation intervals using a joint probability distribution because onset of symptoms and transmission potential jointly depend on the life history of a disease; for example, if an infected individual can only transmit the disease after symptom onset, the forward generation interval will necessarily be longer than the forward incubation period.

The total density of serial intervals between time *p* and *s* can now be obtained by integrating over all possible infection times for the infector and the infectee:

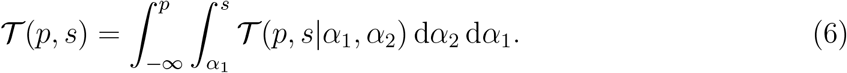

Then, the forward serial-interval distribution *f*_*p*_(*τ*) is given by the density of intervals of length *τ* starting at time *p*, relative to the total number of serial intervals starting at time *p*:

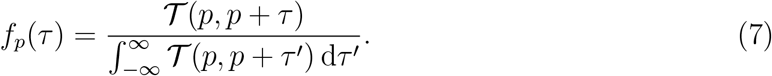

Likewise, the backward serial-interval distribution *b*_*s*_(*τ*) is given by the density of intervals of length *τ* ending at *s*, relative to the total number of serial intervals ending at *s*:

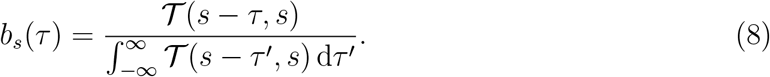

The denominator of the forward serial-interval distribution (Eq. (7)) then corresponds to the total number of infections generated by primary cases who themselves developed symptoms at time *p*. Dividing this quantity by the number of individuals who developed symptoms at time 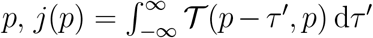, we obtain the serial reproduction number:

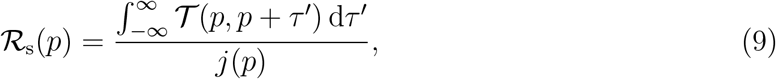

which we define as the average number of infections generated by an individual who developed symptoms at time *p*. Combining the forward serial-interval distribution with the serial reproduction number completes the renewal process between symptomatic cases:

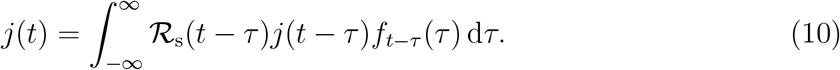

This framework allows us to understand changes in the realized serial intervals for any epidemic model and properly link serial-interval distributions with the renewal process. In addition, assuming that the reproduction number as well as the forward serial-interval distribution remain constant during the exponential growth phase, we can substitute *j*(*t*) ≈ *j*(0) exp(*rt*), ℛ_s_(*t*) ≈ ℛ_s_(0), and *f*_*t*−*τ*_ (*τ*) ≈ *f*_0_(*τ*) to obtain:

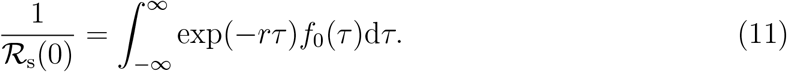

Therefore, the initial forward serial-interval distribution, *f*_0_(*τ*), provides the correct link between the exponential growth rate *r* and the initial serial reproduction number ℛ_s_(0). We re-visit this idea later in Section 2.4 and show that the initial forward serial-interval distribution provides the same *r*–ℛ link as the intrinsic generation-interval distribution.

### 2.3 Epidemic model

We illustrate changes in forward and backward serial intervals over the course of an epidemic by applying our framework to a specific example of an epidemic model. We model disease spread with a renewal-equation model (Heesterbeek and Dietz, 1996; Diekmann and Heesterbeek, 2000; Roberts, 2004; Aldis and Roberts, 2005; Roberts and Heesterbeek, 2007; Champredon et al., 2018). Ignoring births and deaths, changes in the proportion of susceptible individuals *S*(*t*) and incidence of infection *i*(*t*) can be described as:

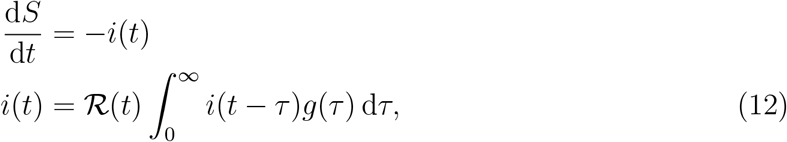

where ℛ (*t*) is the instantaneous reproduction number (i.e., the average number of secondary cases that a primary case infected at time *t* will generate if conditions at time *t* remain unchanged (Fraser, 2007)), and *g*(*τ*) is the intrinsic generation-interval distribution (i.e., the forward generation-interval distribution of a primary case in a population where changes in ℛ(*t*) is negligible (Champredon and Dushoff, 2015)). This model assumes that *g*(*τ*) remains constant through time – in other words, that epidemic dynamics are driven by changes in transmission rate. This assumption may not be well suited to individual-based intervention such as case isolation (Fraser, 2007); nonetheless, this form has been widely used in the literature and has been successfully applied in modeling the current COVID-19 pandemic (Gostic et al., 2020).

Here, changes in reproduction number can be modeled as a product of the basic reproduction number ℛ_0_, proportion susceptible *S*(*t*), and a time-dependent factor *M* (*t*) (for example, accounting for nonpharmaceutical interventions and behavioral changes): ℛ (*t*) = ℛ_0_*S*(*t*)*M* (*t*); Flaxman et al. (2020) used a similar framework to evaluate the impact of nonpharmaceutical interventions on the spread of COVID-19 in 11 countries. Then, the forward generation-interval for a cohort of individuals that were infected at time *p* follows (Champredon and Dushoff, 2015):

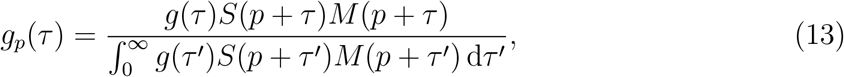

which allows us to separate the joint probability distribution *h*_*p*_ of the forward incubation period and the forward generation-interval distribution as a product of the proportion of susceptible individuals *S* and the joint probability distribution *h* of the forward incubation period and the intrinsic generation intervals:

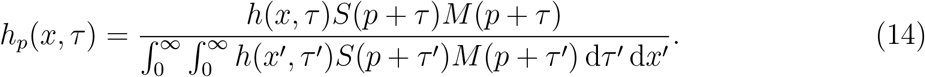

We further assume that the forward incubation period distribution does not vary across cohorts over the course of an epidemic, as it represents the life history of a disease; we denote it as *ℓ*. Then, we have:

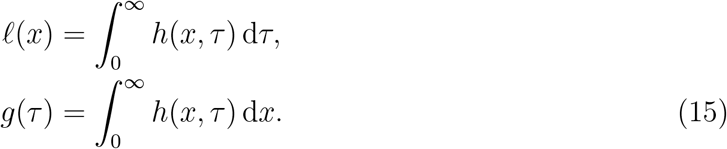

Finally, the case reproduction for this model is defined as follows:

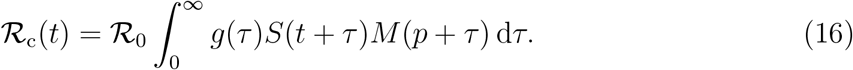

The forward and backward serial-interval distributions are then calculated by substituting these quantities into Eq. (7) and Eq. (8). We use this framework to illustrate how the realized epidemiological time distributions vary over the course of an epidemic and depend on the perspective (i.e., forward vs. backward).

For simplicity, we let *M* = 1 and assume that epidemic dynamics depend only on susceptible depletion in our simulations. Since we are interested in the initial epidemic growth phase (i.e., linking *r* to ℛ), we expect ℛ (*t*) to remain roughly constant during this period. In addition, qualitative effects of *M* that reduces ℛ (*t*) monotonically over time will be similar to the impact of susceptible depletion under this modeling framework. Therefore, general conclusions we draw from our analysis is expected to be robust—however, detailed shape of the epidemic curve and changes in generation- and serial-intervals can still depend on the shape of *M*.

### 2.4 Linking *r* and ℛ

During the initial phase of an epidemic, the proportion susceptible remains approximately constant (*S*(*t*) ≈ *S*(0)) and incidence of infection grows exponentially: *i*(*t*) ≈ *i*_0_ exp(*rt*). During this period, the intrinsic generation-interval distribution provides the correct link between the exponential growth rate *r* and the initial reproduction number ℛ= ℛ_0_*S*(0) based on the Euler-Lotka equation (Wallinga and Lipsitch, 2007). Here, we focus on the estimates of the basic reproduction number *R*_0_ (the value of *R* in a fully susceptible population, *S*(*t*) ≈ 1):

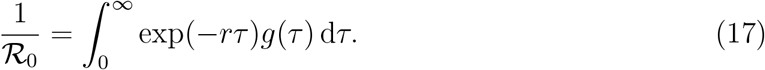

Analogous to the intrinsic generation-interval distribution, forward serial-interval distributions describe the renewal process between symptomatic cases. Therefore, we expect the forward serial-interval distribution during the exponential growth phase — which we refer to as the *initial* forward serial-interval distribution *f*_0_ — to estimate the same value of ℛ_0_ for a given *r* as the intrinsic generation-interval distribution (note, however, that the forward serial interval is not necessarily positive):

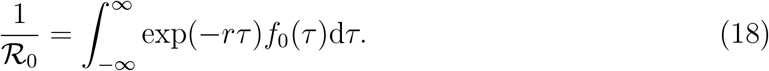

Here, the initial forward serial-interval distribution is given by:

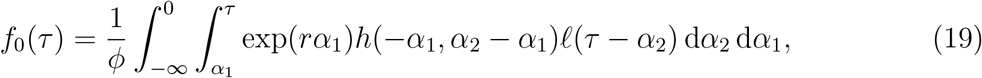

where the normalization constant *ϕ* is determined by the requirement that 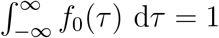. We provide a mathematical proof of this relationship in Supplementary Materials. Since we do not make any assumptions about the shape of the joint distribution *h* between incubation periods and the generation intervals, Eq. (18) holds in general whether or not there is a presymptomatic transmission period.

We further compare this with the estimate of ℛ_0_ based on the intrinsic serial-interval distribution *q*(*τ*):

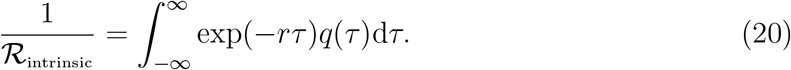

The intrinsic serial-interval distribution *q*(*τ*) does not depend on epidemic dynamics, and is given by:

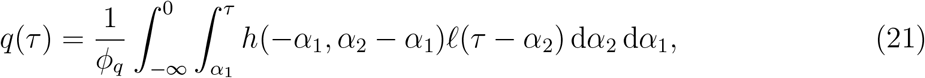

where the normalization constant *ϕ*_*q*_ is determined by the requirement that 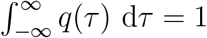. Rather than numerically integrating over closed forms of *g, f*_0_, and *q* to estimate ℛ_0_, we use simulation-based approaches for simplicity (Supplementary Materials).

The initial forward serial-interval distribution depends on the exponential growth rate *r*. For a fast-growing epidemic (high *r*), we expect the backward incubation periods to be short (Eq. (3)), meaning that presymptomatic transmission is less likely to occur. Therefore, the initial forward serial-interval distribution will generally have a larger mean than the intrinsic generation- and serial-interval distributions. However, the exact shape of the initial forward serial-interval distribution depends on the shape of the joint distribution. For example, the Susceptible-Exposed-Infected-Recovered model, under the additional assumption that the incubation and exposed periods are equivalent (i.e. that onset of symptoms and infectiousness occur simultaneously), provides a special case. In this case, the forward serial- and generation-intervals follow the same distributions during the exponential growth phase because (i) infected individuals can only transmit after symptom onset and (ii) the time between symptom onset and infection is independent of the incubation period of an infector (see Supplementary Materials). Everywhere else in this paper, however, we do not assume that the incubation and exposed periods are equivalent. Instead, we allow for presymptomatic transmission in the model in order to reflect the transmission dynamics of COVID-19.

### 2.5 Model parameterization

We have shown that the dynamics of the serial-interval distribution depend on the joint distribution between incubation periods and generation intervals. Here, we use a bivariate lognormal distribution to model the joint probability distribution *h* of intrinsic incubation periods and intrinsic generation intervals (in the renewal equation model, Eq. (12)) while allowing for the possibility that they might be correlated. Given that the viral load of SARS-CoV-2 peaks around the time of symptom onset (He et al., 2020), we generally expect the generation intervals to be positively correlated with the incubation period: that is, individuals who develop symptoms later are more likely to transmit later. Marginal distributions of incubation periods and generation intervals are parameterized based on parameter estimates for COVID-19 (Table 1). For simplicity, we consider four values for the correlation coefficients (on the log scale) of the bivariate lognormal distribution: *ρ* = 0, 0.25, 0.5, 0.75. This parameterization allows for generation intervals to be shorter than the incubation period, allowing for presymptomatic transmission.

**Table 1:**
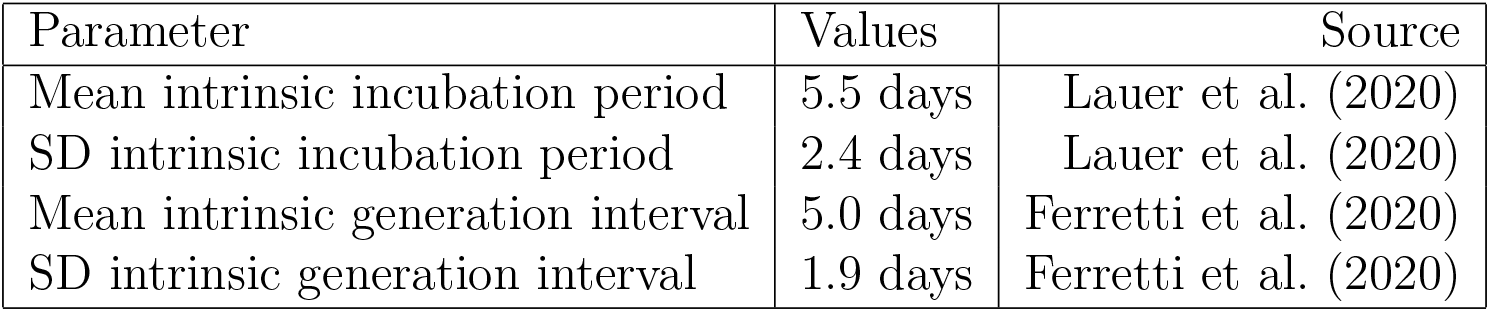
Parameter values used for simulations. The intrinsic incubation period distribution is parameterized using a log-normal distribution with log mean *µ*_*I*_ = 1.62 and log standard deviation *σ*_*I*_ = 0.42. The intrinsic generation-interval distribution is parameterized using a log-normal distribution with log mean *µ*_*G*_ = 1.54 and log standard deviation *σ*_*G*_ = 0.37. Log mean and log standard deviations represent the mean and standard deviations of the underlying normal distributions, which are later exponentiated. The joint probability distribution is modeled using a bivariate log-normal distribution with correlations (on the log scale) *ρ* = {0, 0.25, 0.5, 0.75}. The intrinsic incubation period and generation-interval distributions are chosen to match characteristic of COVID-19 to illustrate realistic magnitudes of time-varying/perspective effects in the current pandemic.

## 3 Results

We use parameter estimates for COVID-19 to characterize the degree to which the realized serial-interval distribution can change over the course of an epidemic and to evaluate how different definitions of the serial-interval distribution can affect the Euler-Lotka estimates of ℛ_0_. We further address how the observed serial intervals, measured through contact tracing, are affected by right censoring during an ongoing epidemic and provide a heuristic method for addressing biases that can arise from using serial-interval data to estimate ℛ_0_. Finally, we analyze serial-interval data from the COVID-19 epidemic in China, outside Hubei province, based on 468 transmission events reported between January 21–February 8, 2020, under our framework.

### 3.1 Realized serial-interval distributions during the exponential growth phase

Fig. 2 shows Euler-Lotka estimates of ℛ_0_ based on different definitions of the serial interval. When the initial forward serial-interval distribution *f*_0_(*τ*) is used, estimates (from Eq. (18)) exactly match the (correct) generation-interval-based estimates (Eq. (17)) for all values of the correlation *ρ* between the intrinsic incubation period and the intrinsic generation interval (Fig. 2A). When the intrinsic distributions are used, however, estimates based on the serial interval (Eq. (20)) underestimate ℛ_0_: as *r* increases, ℛ _intrinsic_ saturates and eventually *decreases* due to the increasing inferred importance of negative serial intervals (Fig. 2B). While the initial forward serial intervals during the exponential growth phase can also be negative, their effects are appropriately balanced because faster epidemic growth leads to longer serial intervals (and a corresponding lower proportion of negative intervals).

**Figure 2:**
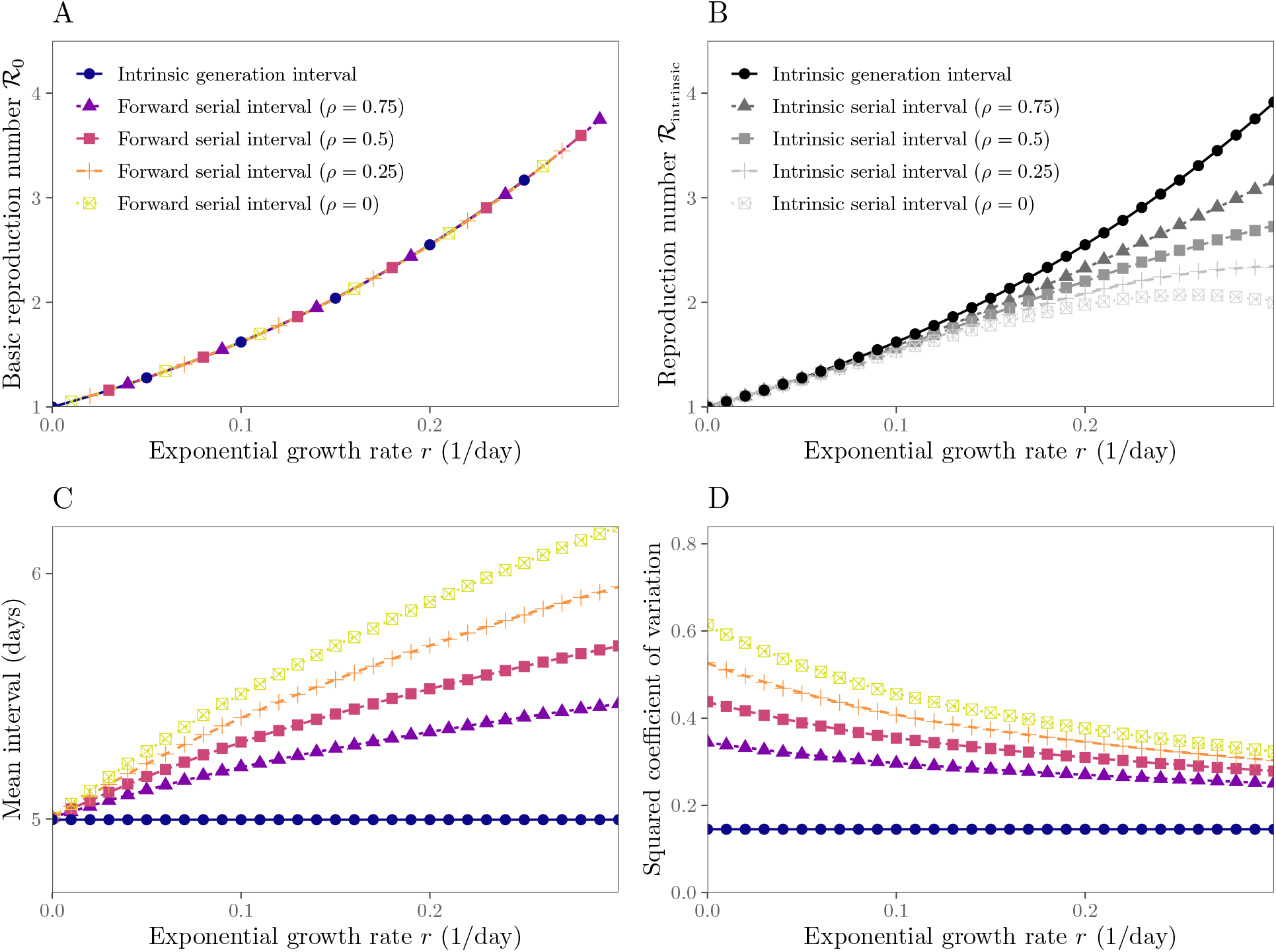
Estimates of the reproduction number from the exponential growth rate based on serial- and generation-interval distributions. (A). The initial forward serial-interval distributions give the correct link between the exponential growth rate *r* and the reproduction number ℛ_0_, for any correlation *ρ* between intrinsic incubation period and intrinsic generation interval of the underlying bivariate log-normal distribution. (B) The intrinsic serial-interval distributions give an incorrect link between *r* and ℛ_0_. (C) The mean initial forward serial interval during the exponential growth phase increases with *r*. (D) The squared coefficient of variation of the initial forward serial intervals during the exponential growth phase decreases with *r*.

Comparing the shapes of the initial forward serial-interval distribution (Eq. (19)) and the intrinsic generation-interval distribution allows us to better understand how different forward distributions lead to identical estimates of ℛ_0_. In general, distributions with higher means and less variability lead to higher ℛ_0_ for a given *r* (Wallinga and Lipsitch, 2007; Weitz and Dushoff, 2015; Park et al., 2019). When incidence is growing exponentially, forward serial intervals have higher means (Fig. 2C) and squared coefficients of variation (Fig. 2D) than the intrinsic generation-interval distribution. The effects of higher means (which increase ℛ_0_) exactly cancel those of higher variability (which decrease ℛ_0_). On the other hand, *intrinsic* serial intervals (Eq. (21)) have the same mean (equal to the mean initial forward serial at *r* = 0 in Fig. 2C) as the intrinsic generation intervals but are more variable (also see squared coefficient of variation of the initial forward serial-interval distribution at *r* = 0 in Fig. 2D); therefore, we underestimate ℛ_0_ when we use the intrinsic serial-interval distribution.

### 3.2 Realized serial-interval distributions during an ongoing epidemic

The initial forward serial-interval distribution captures the exponential growth phase of an epidemic. We now explore how forward and backward serial intervals can vary over the course of an epidemic using deterministic and stochastic simulations based on the renewal equations (see Supplementary Materials) using parameters in Table 1; we further assume ℛ_0_ = 2.5 to reflect the transmission dynamics of COVID-19 in China (Park et al., 2020). While the forward serial-interval distribution is our primary focus, understanding the differences between the forward and the backward distributions is important because the observed intervals during an ongoing epidemic are often the backward ones: we typically identify infected individuals and ask when and by whom they were infected. Similarly, when we are estimating the incubation period of an individual, we typically observe their symptom onset date and try to estimate when they were infected (e.g., Backer et al. (2020)).

Fig. 3 shows the epidemiological dynamics (A) together with the mean forward (B–D) and the mean backward (E–G) delay distributions of a deterministic model based on the renewal equation (Eq. (12)) and of the corresponding stochastic realizations based on individual-based simulations. The mean forward incubation period remains constant throughout an epidemic by assumption (Fig. 3B). The mean forward generation interval decreases slightly when incidence is high, which is when the susceptible population declines rapidly (Fig. 3C; Kenah et al. (2008); Champredon and Dushoff (2015)). In contrast, the mean forward serial interval decreases over time (Fig. 3D).

**Figure 3:**
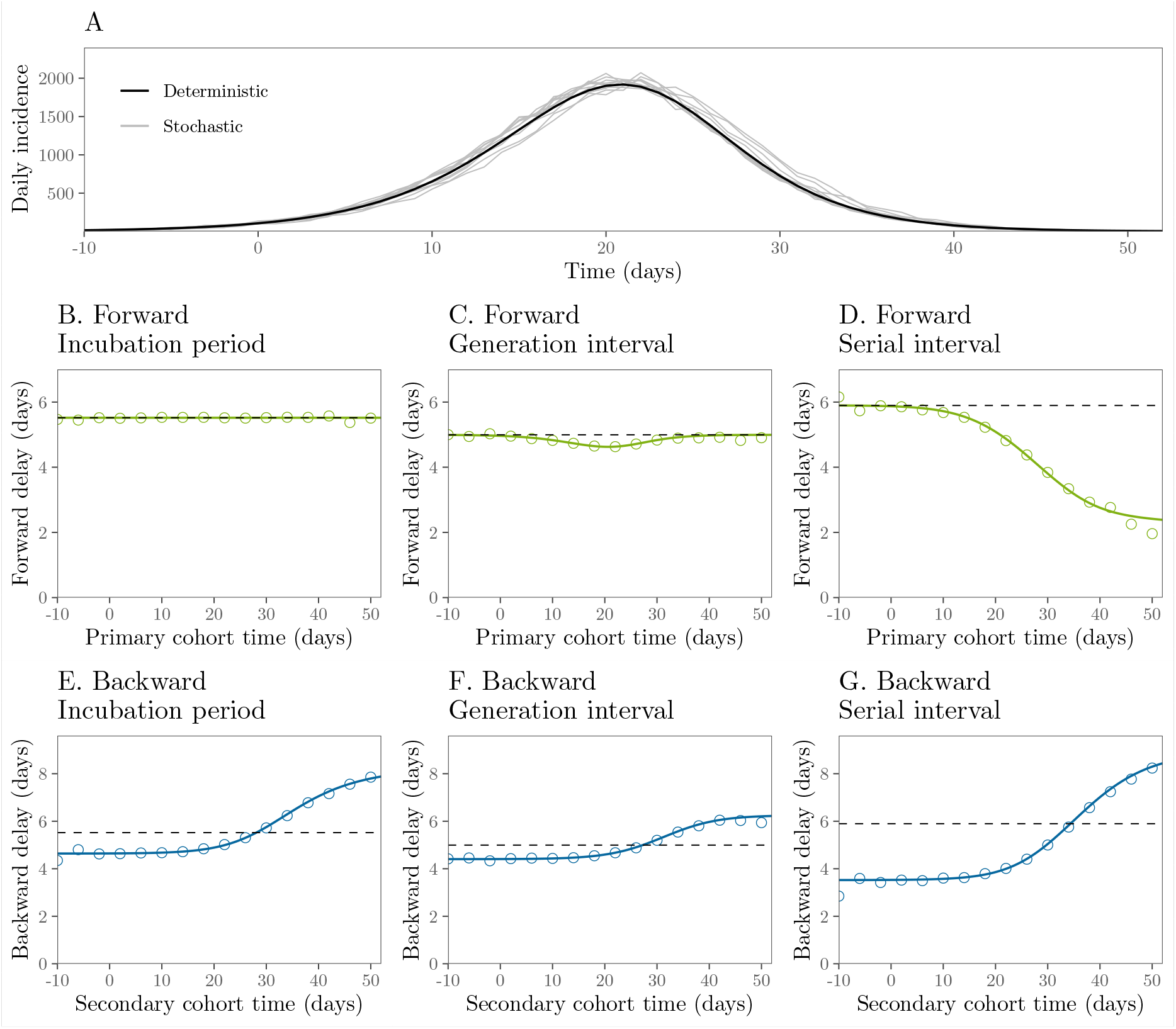
Epidemiological dynamics and changes in mean forward and backward delay distributions. (A) Daily incidence over time. (B–D) Changes in the mean forward incubation period, generation interval, and serial interval. (E–G) Changes in the mean backward incubation period, generation interval, and serial interval. Black (A) and colored (B–G) lines represent the results of a deterministic simulation. Gray lines (A) represent the results of 10 stochastic simulations. Colored points (B-G) represent the average of 10 stochastic simulations. Dashed lines represent the mean initial forward delay. Forward and backward delays are colored according to Fig. 1. In order to remove possible transient dynamics (e.g., left-censoring of time delays and initial stochasticity due to low number of infections), we set *t* = 0 to the first time point when daily incidence is greater than 100. Intrinsic incubation periods and intrinsic generation intervals are assumed to be independent of each other for simplicity. See Supplementary Materials for simulations with correlated incubation periods and generation intervals. See Table 1 for parameter values.

The forward serial-interval distributions depend on distributions of three intervals (Fig. 1B): (i) the backward incubation period, (ii) the forward generation interval, and (iii) the forward incubation period. In these simulations, both forward incubation period (Fig. 3B) and generation-interval (Fig. 3C) distributions remain roughly constant; therefore, changes in the forward serial-interval distributions (Fig. 3D) are predominantly driven by changes in the backward incubation period distribution, whose mean increases over time as the growth rate of disease incidence slows and then reverses. In general, relative contributions of the three distributions depend on their shapes, correlations between intrinsic incubation periods and generation intervals, and overall epidemiological dynamics.

We see similar qualitative patterns in all three backward delays (Fig. 3E–G; Eq. (2)), because they are predominantly driven by the rate of change in incidence, which in turn affects relative cohort sizes. When incidence is increasing, individuals are more likely to have been infected recently, and therefore we are more likely to observe shorter intervals (Eq. (3)). Similarly, when incidence decreases, we are more likely to observe longer intervals. Neglecting these changes will bias the inference of intrinsic distributions from observed distributions.

### 3.3 Observed serial-interval distributions

Now, we turn to practical issues of estimating the reproduction number from the observed serial-interval data during on ongoing epidemic. In order to have an unbiased estimate of the basic reproduction number, we need to estimate the initial forward serial-interval distribution — i.e., serial intervals based on cohorts of infectors who share the same symptom onset time, at the early stage of the epidemic. However, researchers typically use all available information to estimate epidemiological parameters (e.g., aggregating all serial intervals observed until certain time of an epidemic). For example, Thompson et al. (2019) recently suggested that up-to-date serial-interval data are necessary to accurately estimate the reproduction number. We explore the consequences of neglecting changes in the realized serial-interval distribution on estimates of the basic reproduction number.

When an epidemic is ongoing, the observed serial intervals are subject to right-censoring because we cannot observe a serial interval if either an infector or an infectee has not yet developed symptoms. For example, if we were to measure serial intervals on Day 8 as in Fig. 4A, we will only be able to observe the first 6 events (ID 1–6). Fig. 4B demonstrates how the effect of right-censoring in the observed serial intervals translates to the underestimation of the basic reproduction number ℛ_0_ in our stochastic simulations (assuming ℛ_0_ = 2.5 as in Fig. 3). Notably, even if we could observe and aggregate *all* serial intervals across all transmission pairs after the epidemic has ended, we would still underestimate the initial mean forward serial interval (and therefore ℛ_0_), likely by a large amount. The observed serial-interval distribution converges to the intrinsic serial-interval distribution as the incubation periods and generation intervals will no longer be subject to backward biases. In fact, we would even underestimate the intrinsic value slightly due to contraction of the forward generation-interval distribution during the susceptible depletion phase (Fig. 3C). Therefore, aggregated distributions of serial intervals that have been collected throughout different periods of an epidemic must be interpreted with care.

**Figure 4:**
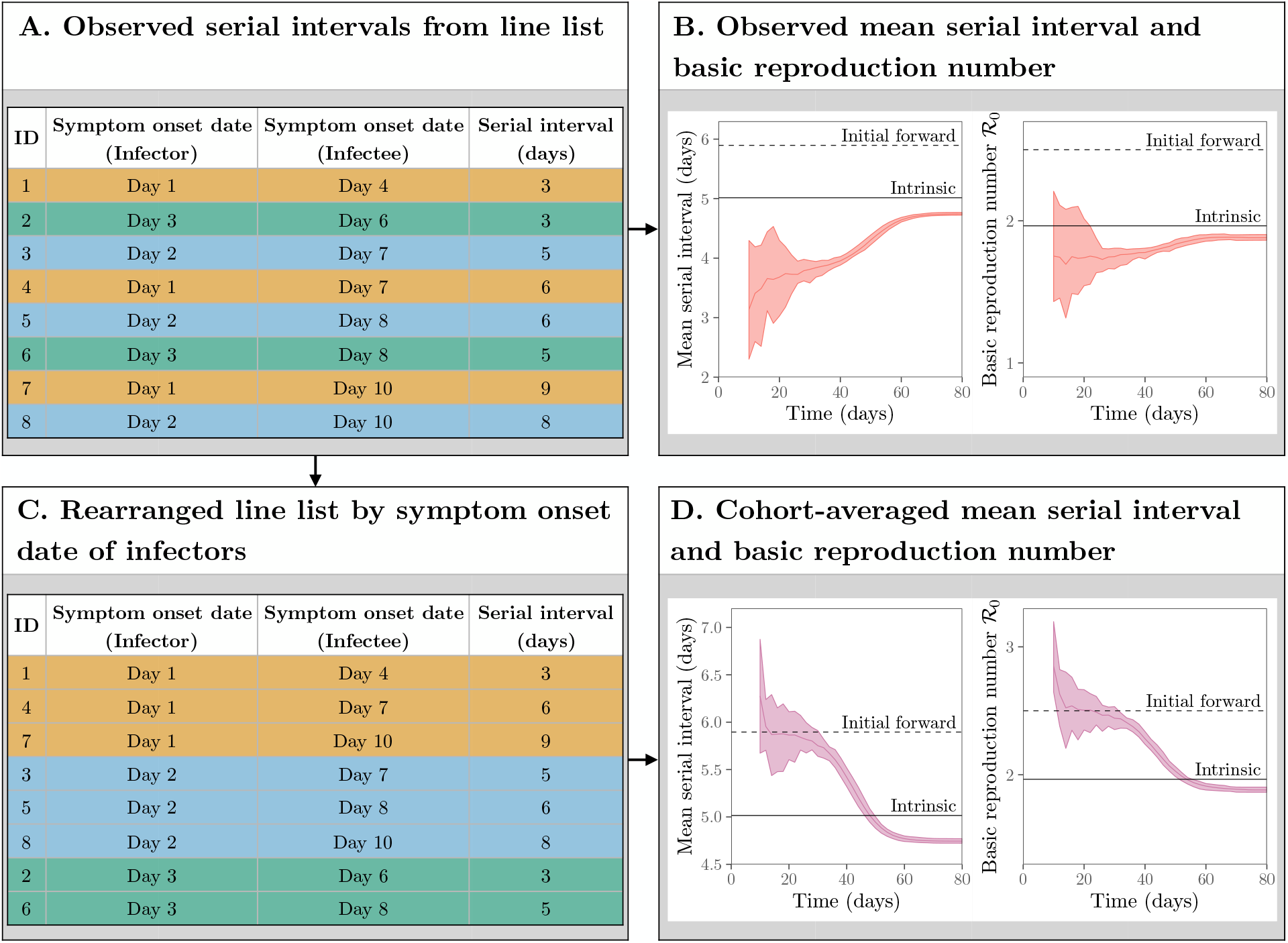
Estimating the reproduction number from the observed serial intervals. (A) Schematic representation of line list data collected during an epidemic. (B) Estimates of ℛ_0_ based on all observed serial intervals completed by a given time. (C) Schematic representation of line list data rearranged by symptom onset date of infectors. (D) Estimates of ℛ_0_ based on all observed serial intervals started by a given time. Black dashed lines represent the mean initial forward serial interval and ℛ_0_. Black solid lines represent the mean intrinsic serial interval and ℛ_intrinsic_. Colored solid lines represent the mean estimates of ℛ_0_ across 10 stochastic simulations. Colored ribbons represent the range of estimates of ℛ_0_ across 10 stochastic simulations.

Here, we provide a heuristic way of assessing potential biases in the estimate of the mean initial forward serial interval and therefore ℛ_0_ retrospectively. We can rearrange the line list and group observed serial intervals based on the symptom onset date of infectors (Fig. 4C)—as we showed earlier, serial intervals that share the same symptom onset date of a primary case give us the forward serial-interval distribution. Then, we can compare how the shape of the serial-interval distribution (particularly its mean) as well as the estimate of ℛ_0_ change as we incorporate more recent cohorts into the analysis: that is, we analyze observed serial intervals from infectors who became symptomatic before time *t* and evaluate how the estimates change as we increase *t*. This approach is analogous to averaging over a set of forward intervals, just as using all information up to a certain time is analogous to averaging over a set of backward intervals (Fig. 4D); the major difference is that we we focus on serial intervals that begin in a certain period, rather than those that end in a certain period. During the exponential growth phase, the estimates of the mean serial interval and ℛ_0_ are consistent with the true value (see ‘initial forward’ in Fig. 4B,D); adding more data allows us to make more precise inference during this period. However, the cohort-averaged estimates decrease rapidly soon after the exponential growth period, reflecting changes in the forward serial-interval distributions. This approach allows us to detect dynamical changes in the forward serial-interval distributions and their effect on the estimates of ℛ_0_.

### 3.4 Applications to the COVID-19 pandemic

Finally, we re-analyze serial intervals of COVID-19 collected by Du et al. (2020) from mainland China, outside Hubei province, based on 468 transmission events reported between January 21–February 8, 2020. Du et al. (2020) estimated the mean serial interval of 3.96 days (95% CI 3.53–4.39 days) and *R*_0_ of 1.32 (95% CI 1.16–1.48). Fig. 5A shows the distribution of symptom onset dates of all individuals within 468 transmission pairs (consisting a total of 752 unique individuals), resembling a COVID-19 epidemic curve in China (cf. Fig. 1 in Pan et al. (2020)). In order to quantify changes in serial intervals, we group them by the symptom onset dates of the primary (Fig. 5B) and secondary (Fig. 5C) cases—corresponding to forward and backward serial-interval distributions, respectively—and compute their mean and 95% quantiles. Fig. 5B shows that the mean forward serial interval decreases over time. While the decrease is likely to be affected by the right-censoring (indicated by the closeness between the quantiles of the observed serial intervals and maximum observable serial intervals), the increase in the proportion of negative serial intervals indicates changes in the forward serial-interval distribution; this proportion is unlikely to be affected by left-censoring (based on the gap between the quantiles of the observed serial intervals and minimum observable serial intervals). The decrease in the mean forward serial interval was probably driven by interventions against spread. Interventions during this time period both decreased (and then reversed) the growth rate of COVID-19 cases — thus increasing the backward incubation period — and also reduced generation intervals, by preventing infections once cases were identified. Both of these would have acted to reduce the forward serial interval. Fig. 5C shows that the mean backward serial interval increased over time, also likely driven directly by the decrease in COVID-19 infections.

**Figure 5:**
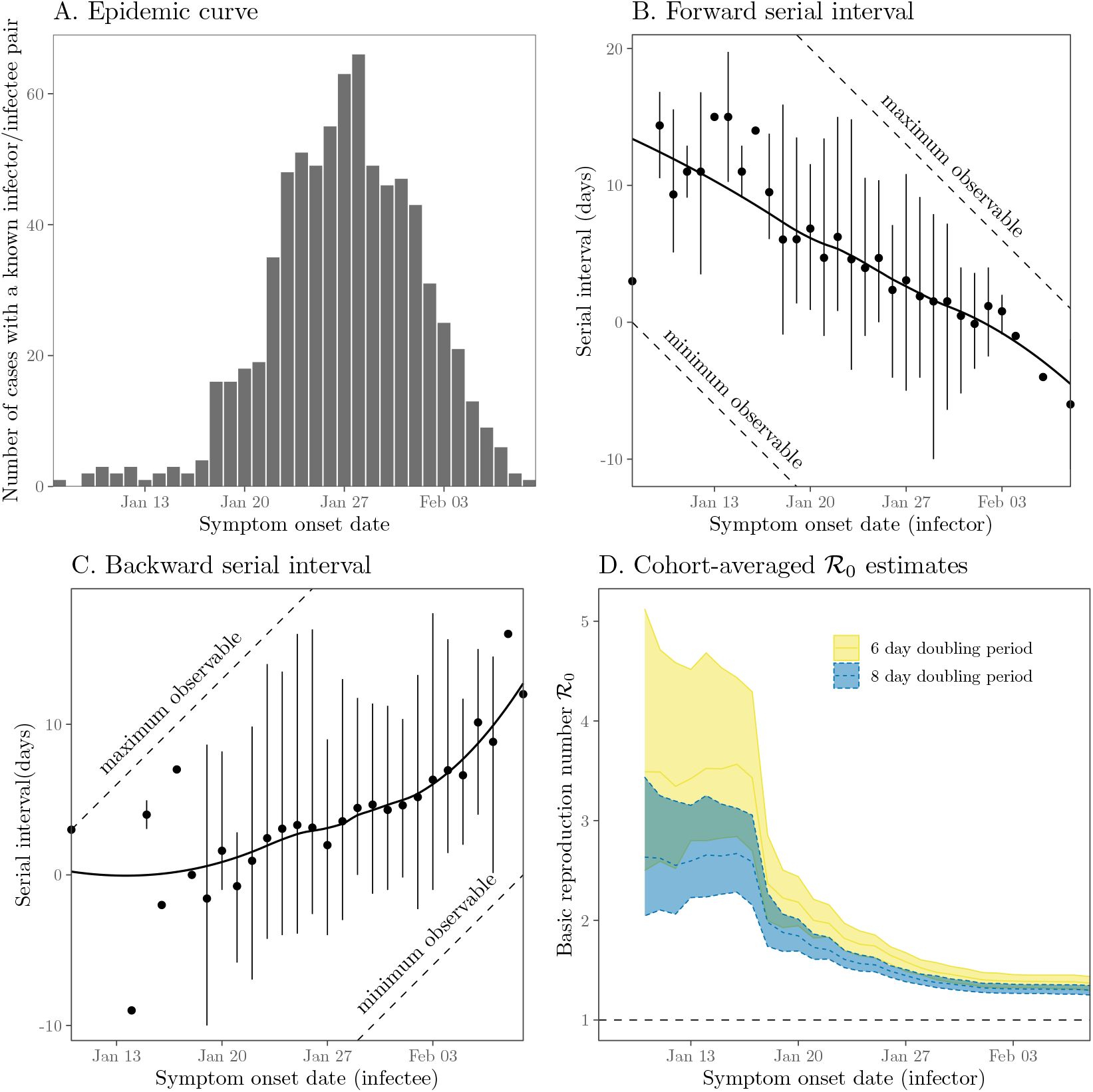
Observed serial intervals of COVID-19 and cohort-averaged estimates of ℛ. (A) Symptom onset dates of all individuals within 468 transmission pairs included in the contact tracing data. (B–C) forward and backward serial intervals over time. Serial interval data have been grouped based on the symptom onset dates of primary (B) and secondary (C) cases. Points represent the means. Vertical error bars represent the 95% equitailed quantiles. Solid lines represent the estimated locally estimated scatterplot smoothing (LOESS) fits. The dashed line represents the maximum and minimum observable delays across the range of reported symptom onset dates. (D) Cohort-averaged estimates of ℛ_0_ assuming doubling period of 6 and 8 days (Li et al., 2020; Wu et al., 2020). Ribbons represent the associated 95% bootstrap confidence intervals. The data were taken from Supplementary Materials of Du et al. (2020).

While the qualitative changes in the mean forward and backward serial interval are consistent with our earlier simulations (Fig. 3), the initial mean forward serial interval (Fig. 5B) appears to be larger than what we calculated based on previously estimated incubation period and generation-interval distributions (Fig. 2C). This difference may imply that the incubation period and generation interval (Table 1) were underestimated, as neither study explicitly accounted for the fact that the observed intervals were drawn from the backward distributions and were likely to have been censored.

Fig. 5D shows the cohort-averaged estimates of ℛ_0_, which remain roughly constant until day January 17th and suddenly decreases; this sudden decrease is due to changes in the forward serial intervals consistent with the dynamics seen in our simulations (Fig. 4). The cohort-averaged estimates of ℛ_0_ based on the early forward serial intervals are also consistent with previous estimates of ℛ_0_ of the COVID-19 epidemic in China (Majumder and Mandl, 2020; Park et al., 2020): ℛ_0_ = 2.6 (95% CI: 2.2–3.1) and ℛ_0_ = 3.4 (95% CI: 2.7 – 4.3) based on a doubling period of 8 or 6 days, respectively, using serial-interval data from infectors who developed symptoms by January 17th. These early cohort-averaged estimates of ℛ_0_ are unlikely to be affected by the right-censoring as we expect the degree of right-censoring to be low (Fig. 5A). Therefore, the original ℛ_0_ estimate of 1.32 (95% CI 1.16-–1.48), which neglects the changes in the forward serial-interval distribution, underestimates ℛ_0_ by a factor of 2.0–2.6. This example demonstrates the danger of using the observed serial intervals to calculate the reproduction number without organizing serial intervals into cohorts.

## 4 Discussion

Generation and serial intervals determine the time scale of disease transmission, and are therefore critical to dynamical modeling of infectious outbreaks. We have shown that the initial *forward* serial-interval distribution — measured from the cohort of infectors who developed symptoms during the exponential growth phase of an epidemic — provides the correct link between the exponential growth rate *r* and the initial reproduction number ℛ. In general, the forward serial-interval distributions will not match the intrinsic serial-interval distribution (which has the same mean as the intrinsic generation-interval distribution) because the incubation period of the infectors (conditional on their symptom onset date of the infector) will be subject to backward biases. In particular, the mean forward serial interval can decrease over time for COVID-19 as individuals who develop symptoms later in an epidemic are more likely to have longer incubation periods, and therefore have greater opportunity to transmit presymptomatically. Failing to account for these effects can result in underestimation of initial ℛ.

Recently, Ali et al. (2020) also showed that forward serial intervals of COVID-19 decreased through time in China. They grouped serial intervals by the symptom onset date of infectors across 14-day periods and found that the mean forward serial interval decreased from 7.8 days to 2.6 days. While they attributed the decrease in serial intervals to reduction of the isolation delay, their regression analysis showed that isolation delays explain only 51.5% of the variation in serial intervals (they could explain up to 72% of the variance by including other intervention measures). Our framework provides an explanation for the remaining variation: changes in the backward incubation period during the decreasing phase of an epidemic act to further shorten serial intervals due to increased amount of presymptomatic transmission (even in the absence of nonpharmaceutical interventions). Isolation delays and other intervention measures affect the amount of onward transmission, and therefore the distribution of realized (forward) generation intervals. They therefore are not expected to explain all the variation in forward serial intervals, since these additionally depend on both the backward incubation period of the infector and the forward incubation period of the infectee (Fig. 1B).

Our results support the use of serial-interval distributions for calculating the ℛ during the exponential growth phase, but they also reveal gaps in current practices in incorporating serial-interval distributions into outbreak analyses. For example, Thompson et al. (2019) recently emphasized the importance of using up-to-date serial-interval data for accurate estimation of time-varying reproduction numbers. However, our results show that if observational biases in the forward serial interval through time are not accounted for, using up-to-date serial-interval data can actually exacerbate the underestimation of ℛ in the initial growth phase of an outbreak. Future studies should explore how neglecting changes in the forward serial-interval distribution can affect the estimates of ℛ beyond the exponential growth phase, and potentially re-assess existing estimates of ℛ. We also suggest that modelers should aim to characterize spatiotemporal variation in forward serial-interval distributions. These modeling approaches should be coupled with epidemiological investigation through contact tracing. Going forward, an additional advantage of early, intensive contact tracing of emerging diseases is that it provides the best information to characterize the initial forward serial-interval distribution.

Our study underlines the fact that the serial-interval distribution depends not only on the generation-interval and incubation-period distributions, but also on the correlation between their duration in a given individual. Here, we use a bivariate lognormal distribution to capture these correlations phenomenologically and to show that realized serial intervals can decrease over time in the context of COVID-19. Although their true correlation will depend on viral load dynamics, we expect our conclusions about decreasing serial intervals of COVID-19 to be robust, as individuals with longer incubation periods will generally have a longer time window to transmit before symptom onset. In general, the impact of increasing backward incubation periods on the forward serial intervals are likely to be disease-specific— for example, we show in Supplementary Materials that the initial forward serial-interval distribution can be equivalent to the intrinsic generation-interval distribution, regardless of the growth rate *r*, due to independence between the incubation period and time from symptom onset to transmission and the lack of presymptomatic transmission. Future studies trying to interpret realized serial intervals should consider carefully the joint distribution between the generation intervals and incubation periods.

In closing, we lay out a few practical principles for analyzing and interpreting serial-interval data. First, serial intervals should be cohorted based on the symptom onset date of the infector (and not of the infectee) whenever possible. Previous studies have often regarded serial intervals as an intrinsic quantity, having the same mean as the intrinsic generation interval (Svensson, 2007; Klinkenberg and Nishiura, 2011; Champredon et al., 2018; Britton and Scalia Tomba, 2019), but the distribution (and the mean) of observed serial intervals differs from this expectation, and changes through time due to epidemic dynamics. Second, aggregating serial intervals across different cohorts and epidemic periods should be avoided because the realized serial-interval distribution can be subject to different censoring and epidemiological biases: Even when *all* realized serial intervals can be observed throughout an unmitigated epidemic, we do not obtain the intrinsic serial interval distribution due to susceptible depletion (Fig. 4). Third, applying serial-interval information across epidemics of a given disease should be done with care, because serial intervals are epidemic-specific, rather than disease-specific. Finally, serial-interval data should be accompanied by a trajectory of the epidemic curve, whenever possible, to provide epidemiological context. In practice, these recommendations will sometimes be hard to follow, due to limited data about serial intervals, but these issues should be kept in mind when interpreting serial-interval data to inform transmission dynamics.

More broadly, our study underlines the importance of carefully defining measured epidemiological time distributions. Previous studies have shown the importance of forward vs. backward measurement of generation intervals (Nishiura, 2010; Champredon and Dushoff, 2015; Britton and Scalia Tomba, 2019); we generalize these ideas and show that they apply to other epidemiological distributions. Some studies during the early phases of the COVID-19 epidemics have tried to correct for the backward biases (Verity et al., 2020), but changes in the backward delay distributions due to changing cohort sizes are expected to be a pervasive feature of outbreak dynamics. Cohorting epidemiological delays by the primary event time can help avoid backward biases (although censoring biases can still exist) as well as detect potential changes in the distribution.

Here, we assume that all individuals develop symptoms and that the entire transmission process, including all relevant epidemiological delays, is known exactly. In practice, identifying who infected whom is difficult in general, and asymptomatic and presymptomatic transmission of COVID-19 exacerbates this difficulty (Bai et al., 2020; He et al., 2020; Wei, 2020). Biases in the observed serial intervals will necessarily bias the estimates of ℛ. Furthermore, when one of the individuals in a transmission pair is asymptomatic, there is no symptom-based serial interval. Neglecting the time scale of asymptomatic transmission may also bias the estimates of ℛ (Park et al., 2020).

Despite these limitations, our analysis of serial intervals of COVID-19 from China provides further support for our theoretical framework, demonstrating temporal variation in serial intervals and its effect on the estimates of ℛ. Most existing estimates of the serial-intervals of COVID-19 implicitly or explicitly assume that the serial-interval distributions remain constant throughout the course of an epidemic (Du et al., 2020; He et al., 2020; Nishiura et al., 2020; Tindale et al., 2020; Zhao et al., 2020; Zhang et al., 2020). Our study provides a rationale for reassessing estimates of serial-interval distributions—and their use in estimating ℛ—during the COVID-19 pandemic.

## Data Availability

All code is available at: https://github.com/parksw3/serial

## Data availability

All data and code are stored in a publicly available GitHub repository (https://github.com/parksw3/serial).

## Competing interests

We declare no competing interests.

## 5 Supplementary Materials

### 5.1 Deterministic simulation

We simulate the renewal equation model using a discrete-time approximation:

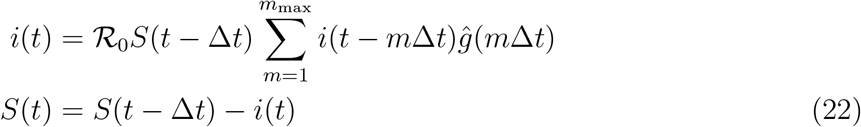

where *ĝ* is a discrete-time intrinsic generation-interval distribution that satisfies the following:

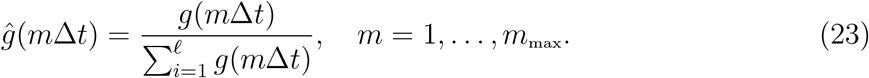

The continuous-time intrinsic generation-interval distribution is parameterized using a log-normal distribution (Table 1). We define the intrinsic incubation period distribution in a similar manner:

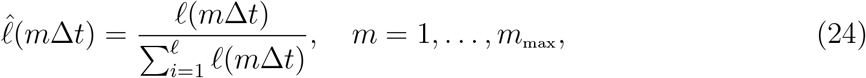

where its continuous-time analog is also based on a log-normal distribution. For simplicity, we assume that the forward incubation periods and intrinsic generation intervals are independent:

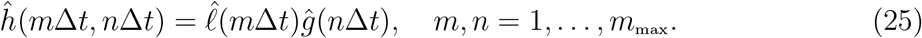

We use Δ*t* = 0.025 days and *m*_max_ = 2001 for discretization steps.

We initialize the simulation with population size *N* =40,000 as follows:

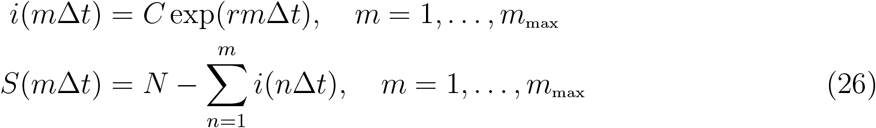

where *C* is chosen such that 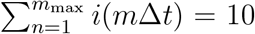. These initial conditions allow the model to follow exponential growth from time Δ*t*(*m*_max_ + 1) without any transient behaviors.

### 5.2 Stochastic simulation

We run stochastic simulations of the renewal equation model using an individual-based model on a fully connected network (i.e., homogeneous population) based on the Gillespie algorithm that we developed earlier (Park et al., 2020). First, we initialize an epidemic with *I*(0) infected individuals (nodes) in a fully connected network of size *N*. For each initially infected individual, we draw number of infectious contacts from a Poisson distribution with the mean of ℛ_0_ and the corresponding generation intervals for each contact from a log-normal distribution (Table 1). Contactees are uniformly sampled from the total population.

All contactees are sorted into event queues based on their infection time. We update the current time to the infection time of the first person in the queue. Then, the first person in the queue makes contacts based on the Poisson offspring distribution described earlier and their contactees are added to the sorted queue. Whenever contactees are added to the sorted queue, we remove all duplicated contacts (but keep the first one) as well as contacts made to individuals that have already been infected. Simulations continue until there are no more individuals in the queue. We simulate 10 epidemics with *I*(0) = 10 and *N* =40,000.

### 5.3 Linking *r* and ℛ_0_ using serial-interval distributions

The intrinsic generation-interval distribution *g*(*τ*) provides a link between *r* and ℛ_0_ via the Euler-Lotka equation (Wallinga and Lipsitch, 2007):

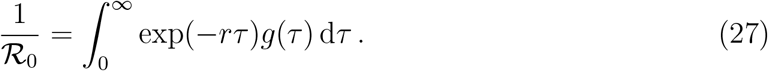

In this section, we prove that the initial forward serial-interval distribution *f*_0_(*τ*) also estimates the same ℛ_0_ from *r*, except that integral extends to *τ* = − ∞ rather than beginning at *τ* = 0, because serial intervals can be negative:

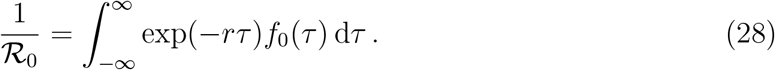

Here, the initial forward serial-interval distribution *f*_0_(*τ*) is defined as:

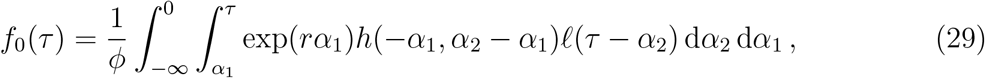

where *h* is the joint probability distribution describing the intrinsic generation-interval distribution *g* and the intrinsic incubation period distribution 𝓁(see Eq. (15) in the main text), and the normalization constant *ϕ* is determined by the requirement that 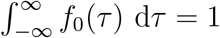.

In order to verify Eq. (28), we first rewrite the integral in Eq. (29) by substituting −*α*_1_ for *α*_1_, and then changing the order of integration:

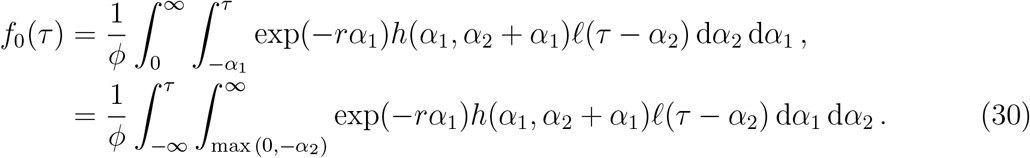

To further simplify the expression, we define *z*(*α*_2_) as follows:

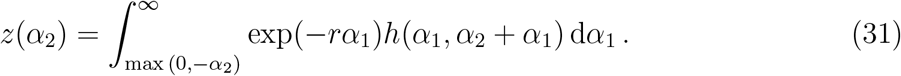

Substituting *z*(*α*_2_) into Eq. (30) we obtain:

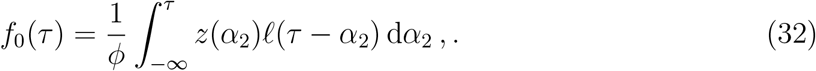

Writing 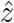 for a normalized version of *z*,

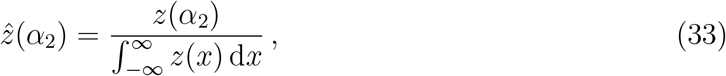

we can now express the initial forward serial-interval distribution *f*_0_ as a convolution of 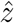 and 𝓁:

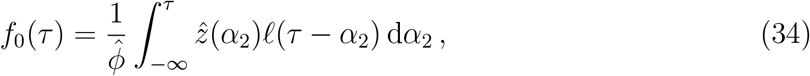

where 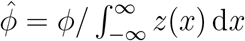.

Since the right hand side of Eq. (28) is also a Laplace transform of 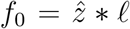, we can express it as the product of Laplace transforms of 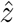 and 𝓁:

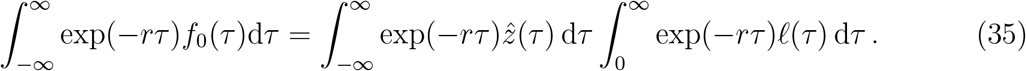

In order to derive an expression for a Laplace transform of 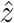, we have to first derive an analytical expression for 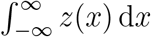. By changing the order of integration, we have:

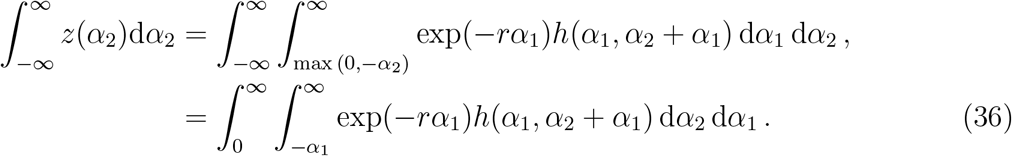

Since 𝓁 is a marginal probability distribution of *h*, it follows that:

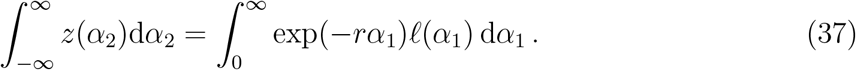

Then, we have:

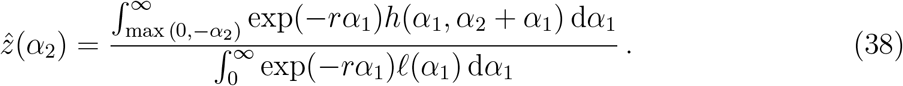

Substituting the expression into Eq. (35), we have:

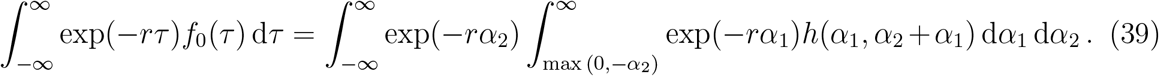

Recall that *g* is also a marginal probability distribution of *h*:

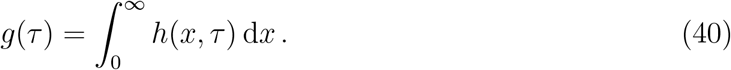

We can then substitute *τ* = *α*_1_ + *α*_2_ into Eq. (39) and apply change of variables to obtain:

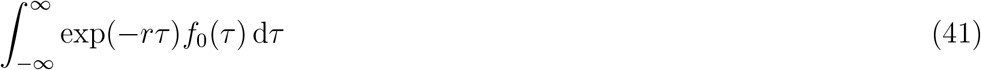

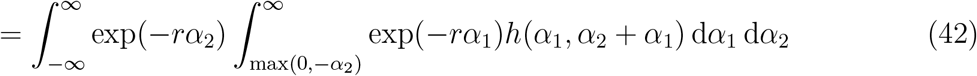

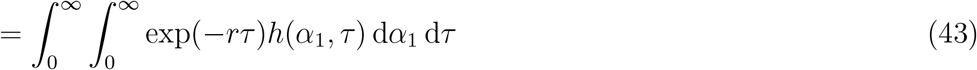

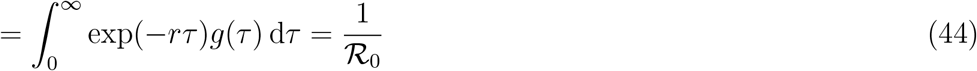

Therefore, the initial forward serial-interval distribution and the intrinsic generation-interval distribution give the same estimates of ℛ_0_ from *r*. □

### 5.4 Comparing the estimates of ℛ_0_ using the initial forward and the intrinsic serial-interval distributions

We use a simulation-based approach to compare the estimates of ℛ_0_ based on the serial- and generation-interval distributions. To do so, we model the intrinsic generation-interval distribution and the incubation period using a multivariate log-normal distribution with log means *µ*_*G*_, *µ*_*I*_, log standard variances 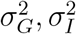, and log-scale correlation *ρ*; the multivariate log-normal distribution is parameterized based on parameter estimates for COVID-19 (Table 1). We construct forward serial intervals during the exponential growth period as follows:

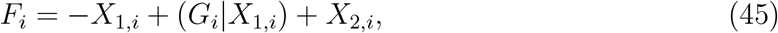

where the backward incubation period *X*_1,*i*_ of an infector is simulated by drawing random log-normal samples *Y*_*i*_ with log mean *µ*_*I*_ and log variance 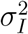 and resampling *Y*_*i*_, each weighted by the inverse of the exponential growth function exp(−*rY*_*i*_); the intrinsic generation interval conditional on the incubation period of the infector (*G*_*i*_|*X*_1,*i*_) is drawn from a log-normal distribution with log mean *µ*_*G*_ + *σ*_*G*_*ρ*(log(*X*_1,*i*_) − *µ*_*I*_)*/σ*_*I*_ and log variance 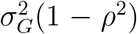; the forward incubation period *X*_2,*i*_ of an infectee is drawn from a log-normal distribution with log mean *µ*_*I*_ and log variance 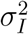. We then calculate the basic reproduction number ℛ_0_ using the empirical estimator:

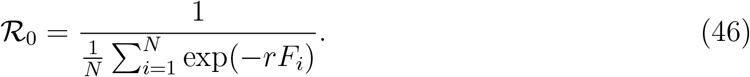

We compare this with an estimate of ℛ_0_ based on the intrinsic serial-interval distribution which has the same mean as the intrinsic generation-interval distribution (Svensson, 2007; Klinkenberg and Nishiura, 2011; Champredon et al., 2018; Britton and Scalia Tomba, 2019):

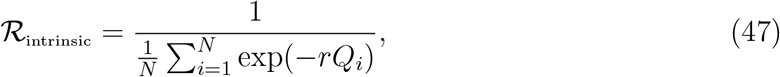

where

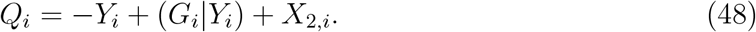

### 5.5 Applications: SEIR model

Consider a Susceptible-Exposed-Infectious-Recovered model:

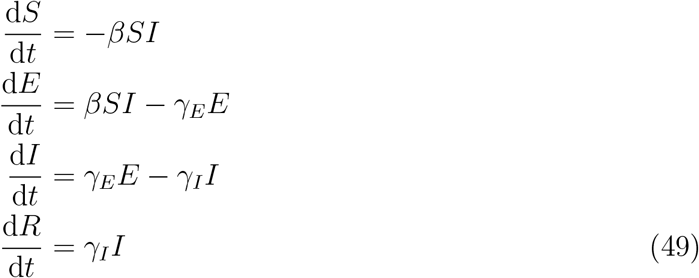

where *β* is the transmission rate, 1*/γ*_*E*_ is the mean latent period, and 1*/γ*_*I*_ is the mean infectious period. We further assume that the latent period is equivalent to incubation period; in other words, infected individuals can only transmit after symptom onset. Then, the generation interval will be always longer than the incubation period.

The joint probability distribution of the intrinsic incubation periods and intrinsic generation intervals for this model can be written as:

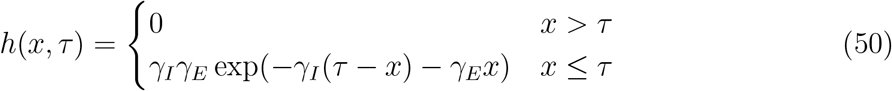

Then, the intrinsic generation-interval distribution is given by:

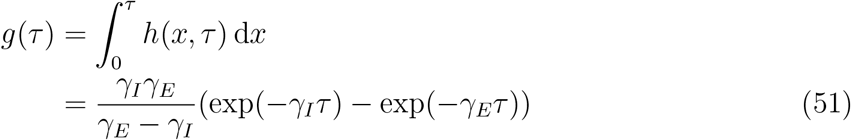

On the other hand, the initial forward serial-interval distribution is given by:

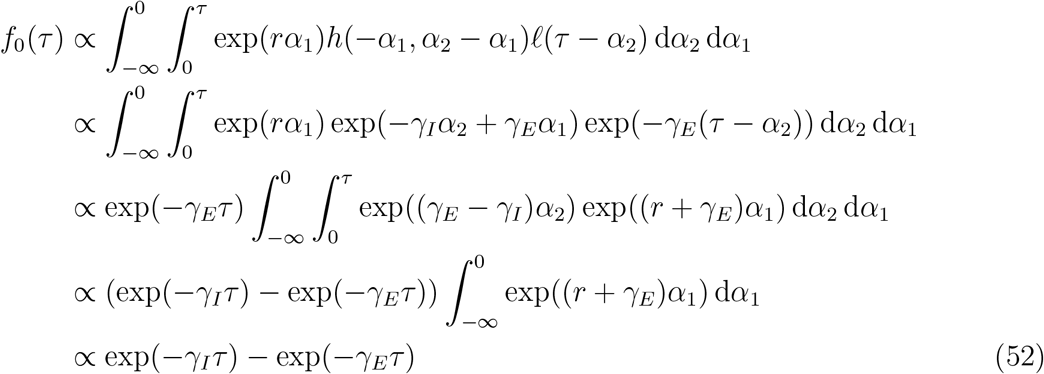

Therefore, both the intrinsic generation intervals and the initial forward serial intervals are identically distributed and have the same mean.

### 5.6 Simulations with correlated intrinsic incubation periods and intrinsic generation intervals

**Figure S1:**
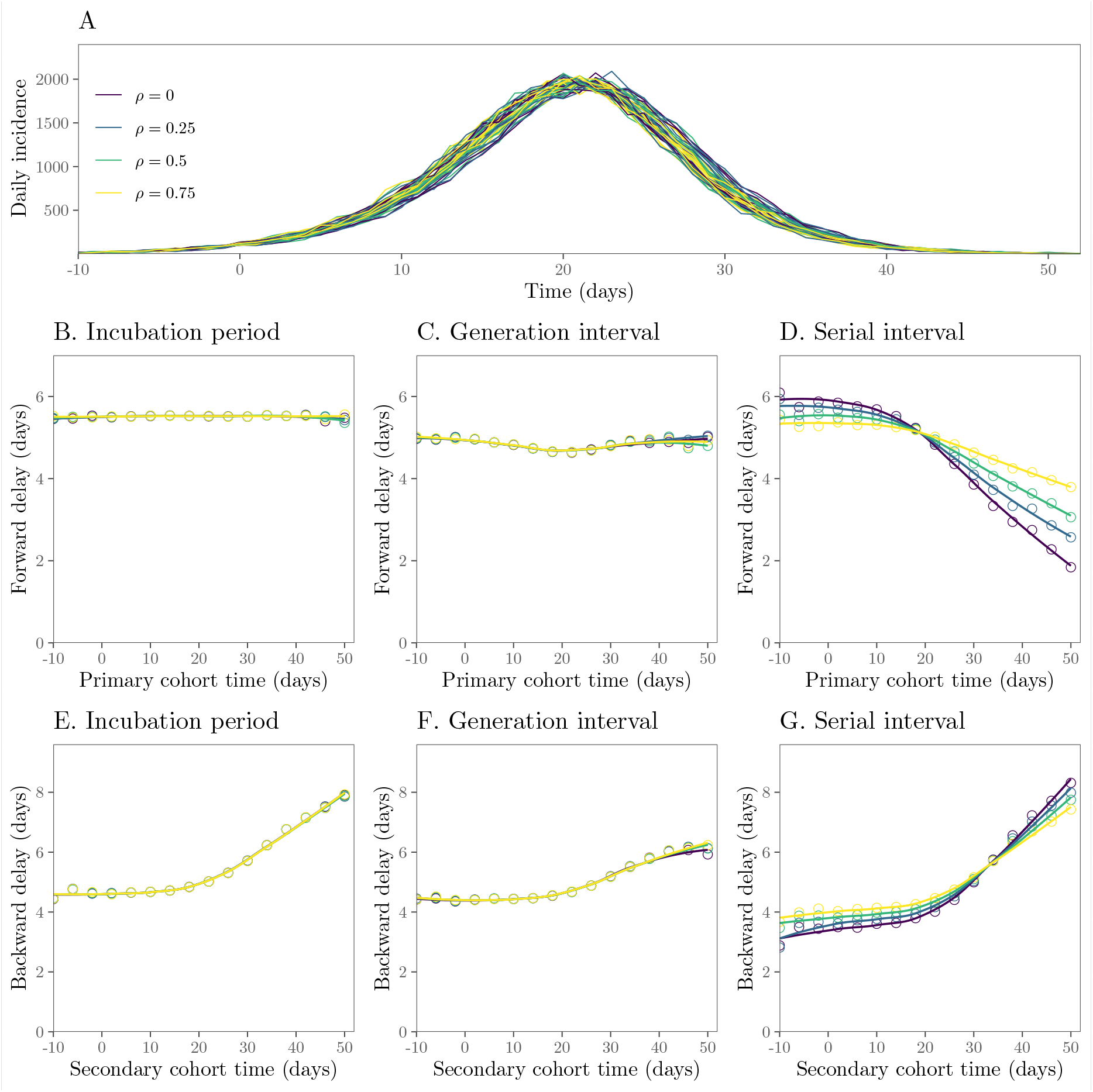
Epidemiological dynamics and changes in mean forward and backward delay distributions. (A) Daily incidence over time. (B–D) Changes in the mean forward incubation period, generation interval, and serial interval. (E–G) Changes in the mean backward incubation period, generation interval, and serial interval. Intrinsic incubation periods and intrinsic generation intervals are modeled using a correlated bivariate lognormal distribution; therefore, generation intervals are drawn from the corresponding conditional distributions (given a incubation period), instead of the marginal distribution. Higher correlation reduces the amount of changes in the mean forward serial interval because shorter (longer) backward incubation periods of infectors during the increasing (decreasing) phase of an epidemic are associated with shorter (longer) forward generation intervals. See Figure 3 in the main text for a detailed description.

## Notes

### Competing Interest Statement

The authors have declared no competing interest.

### Funding Statement

.

### Author Declarations

No IRB approvals were required for the work.

